# Fatigue is a key contributor to quality of life in heart valve disease and after valve replacement/repair

**DOI:** 10.1101/2025.07.11.25331403

**Authors:** Ariel Pons, Gillian Whalley, Rosemary Wyber, Paul Bridgman, Ralph Stewart, Philip Adamson, Ross Roberts-Thompson, Crispin Jenkinson, David Morley, Sean Coffey

## Abstract

**Background:** Heart valve disease can result in high morbidity and impairment of quality of life (QOL) both before and after intervention. However, there are few descriptions of the QOL of people with heart valve disease across the disease course.

**Aims:** We aimed to describe the QOL of people living with heart valve disease through qualitative interviews.

**Methods:** Semi-structured interviews were conducted in people with heart valve disease, their family members, and clinical experts. A simple thematic analysis was used to summarise their perceptions of QOL.

**Results:** We interviewed 34 people with heart valve disease: seven with aortic stenosis, seven with rheumatic heart disease involving the mitral valve, nine with mitral regurgitation, and 11 with valve replacement/repair (mean age 66, 56% female). Three family members and five clinical experts were also interviewed. A key contributor to QOL was fatigue: most participants experienced fatigue, even mild fatigue impaired QOL directly, and severe fatigue had devastating effects on quality of life. Physical limitations impaired QOL due to the loss of ‘normal’ activity rather than objective physical limitation. Symptoms of heart valve disease impaired QOL directly, but the indirect effects of valve disease inspiring worry that reduced confidence and activity led to greater impairment.

**Conclusions:** Fatigue both before and after valve intervention is a contributor to QOL and requires further assessment. Research is recommended into whether fatigue is a specific enough symptom to warrant valvular intervention in heart valve disease.

## INTRODUCTION

With a decreasing mortality rate, there is an increasing prevalence of people living with heart valve disease (HVD) worldwide.[1] This requires an increasing focus on quality of life (QOL) in HVD, and in particular to find ways to improve it. However, to date there has been little formal research into QOL in HVD, most of it using instruments developed for other conditions, especially heart failure.[2] In this study, we wish to describe the components of QOL in HVD from first principles.

We focus on the three most common forms of HVD: rheumatic heart disease (RHD), aortic stenosis (AS), mitral regurgitation (MR), as well as patients with a valve replacement/repair (VRR). Globally, RHD is the commonest form of HVD, with an incidence of nearly 3 million per year.[1] It is due to the sequelae of infection by Group A Streptococci and mostly occurs in young populations in low-income countries. In high-income countries, AS and MR are more common, with AS being caused usually by a degenerative calcification process, and MR occurring in middle and older age due to a wide range of primary and secondary causes. New Zealand and Australia have higher rates of RHD than in comparable countries,[1] with Indigenous and socially disadvantaged populations overwhelmingly disproportionately affected.

## METHODS

Ethical approval was granted by New Zealand’s Health and Disabilities Ethics Committee (HDEC, ethics reference 19/NTA/163). All individuals gave informed consent to take part in the study, and participant anonymity was preserved in analysis and presentation. To identify determinants of QOL in HVD, we interviewed people with HVD, family members, and clinical experts. This is the first phase of a four phase process in developing a HVD-specific instrument to quantify QOL in HVD, called the VALVQ, with the full pre-specified protocol available elsewhere.[3]

### Participant identification and recruitment

People with HVD (where we define HVD to mean RHD, AS, MR, and VRR) were identified using the echocardiographic databases in New Zealand’s Southern District Health Board and Canterbury District Health Board between October 1 to December 31 2019. Eligibility was assessed using the electronic patient record. Eligible patients were approached in consecutive order for potential recruitment. Pre-specified target recruitment numbers were 10 participants in each form of HVD (minimum 5), 3-5 family members/carers, and 5 clinical experts. Family members were recruited by asking recruited participants, and clinical experts were identified by the corresponding author (SC).

### Inclusion and exclusion criteria

To be included in this project, individuals had to have clinically significant primary heart valve disease or a repair/replacement of a damaged valve as their primary diagnosis, and awareness of their disease. Patients were excluded if they had ischaemic heart disease or any other heart comorbidity other than well-controlled atrial fibrillation, were younger than 18, had a terminal illness or cognitive/mental impairment, had a severe comorbidity or comorbidity significant enough to be expected to have a greater effect on the person’s QOL than their HVD, had recently been hospitalised, or were pregnant (due to the changes in cardiac and valvular function during pregnancy). These exclusions were based primarily on the electronic health record. Family members of recruited patients in phase one had to be over the age of 18 and have awareness of the patient’s HVD.

### Interviews

Interviews were conducted by investigator AP over a phone call or Zoom meeting, with in-person interviews not possible due to nationwide COVID-19 lockdowns in place at the time. Interviews with HVD participants were semi-structured, with the interview structure being determined by the first few interviews. The initial and the final structures of the interviews are included in the Appendix (Appendix Table One: Interview Structure). Interviews continued until saturation was reached (when no new concepts were emerging).

All interviews were recorded and transcribed (AP).

### Analysis

Data coding was not used. In accordance with our aims of representing patients’ experiences, an iterative thematic analysis was used.[4]

## RESULTS

### Participant demographics

125 eligible patients were identified by the search and invited to participate. 34 (27%) patients with HVD participated. Of the 91 who did not participate, most (80%) could not be initially contacted, 11% declined participation, and 7% were lost to follow-up. Participants were similar age to non-participants (mean age 67 vs 62 years, ns) and no differences in sex distribution were seen (female 53% vs 44%, ns). There were no significant differences between the HVD type, HVD severity, or ethnicity of participants compared to non-participants.

A total of 42 participants were interviewed. Five were clinical experts, three were family members of participants with HVD, and 34 were patients with HVD (Table one): seven with AS, seven with RHD, nine with MR, and 11 with replaced or repaired aortic or mitral valves. Of the 34 HVD participants, mean age was 66 years (min 19, max 93), 56% were female, and 85% were of New Zealand European ethnicity. The clinical experts, recruited from New Zealand and Australia, consisted of a clinical cardiologist, an imaging cardiologist, two structural interventional cardiologists, and a general practitioner with experience of care for rural patients with RHD.

**Table one:** Participant Demographics.

|  | Total sample | Aortic Stenosis | Mitral Regurgitation | Rheumatic Heart Disease | Valve Replacement /Repair |
| --- | --- | --- | --- | --- | --- |
| **n** | 34 | 7 | 9 | 7 | 11 |
| **Mean age, years (SD)** | 66.1 (16.0) | 64.1 (7.6) | 70.3 (8.4) | 47.0 (20.5) | 76.0 (11.0) |
| **Female sex (%)** | 19 (55.9) | 6 (85.7) | 3 (33.3) | 1 (14.3) | 5 (45.5) |
| **Ethnicity (%)** |  |  |  |  |  |
| NZ European | 28 (82.4) | 6 (85.7) | 8 (88.9) | 3 (42.9) | 11 (100) |
| Samoan | 2 (5.9) | 0 | 0 | 2 (28.6) | 0 |
| Māori | 1 (2.9) | 0 | 1 (11.1) | 0 | 0 |
| European | 1 (2.9) | 1 (14.3) | 0 | 0 | 0 |
| Fijian | 1 (2.9) | 0 | 0 | 1 (14.3) | 0 |
| Filipino | 1 (2.9) | 0 0 | 0 | 1 (14.3) | 0 |
| **HVD severity (%)** |  |  |  |  |  |
| Mild | 2 (5.9) | 0 | 0 | 2 (28.6) | NA |
| Moderate | 6 (17.6) | 4 (57.1) | 1 (11.1) | 1 (14.3) | NA |
| Moderate-severe | 6 (17.6) | 1 (14.3) | 3 (33.3) | 2 (28.6) | NA |
| Severe | 7 (20.6) | 2 (28.6) | 5 (55.6) | 0 | NA |
| **Mixed HVD (%)** | 3 (8.8) | 1 (14.3) | 0 | 2 (28.6) | 0 |
| **Respiratory disease (%)** | 3 (8.8) | 2 (28.6) | 1 (11.1) | 0 | 0 |
| **Prior cancer (%)** | 5 (14.7) | 0 | 1 (11.1) | 0 | 4 (36.4) |
| **Systemic inflammatory disease (%)** | 3 (8.8) | 0 | 1 (11.1) | 1 (14.3) | 1 (9.1) |

HVD = Heart Valve Disease; NZ = New Zealand; SD = Standard Deviation.

### Interview Findings

Interviews were conducted between March 2020 and October 2020. Interviews with patients and family members took between 10 and 35 minutes, and Interviews with clinicians took approximately 30 minutes. Interview content was summarised into themes, broadly classified as physical factors, perspective, and stressors. Details of themes with participant quotes are available in the appendix as Appendix Table Two: Physical factors and Quality of Life; and Appendix Table Three: Perspective, Stressors, and Quality of Life.

Physical limitation was an important determinant of QOL: participants often defined their QOL as their physical capacity/limitation. Loss of physical ability not only directly reduced QOL but also prevented participants from completing activities they considered ‘normal’ and a part of their ‘usual’ life; losing this caused an additional sense of loss which further reduced QOL. Shortness of breath (SOB) and pain, both of which are common HVD symptoms, had a similar effect on QOL. As might be expected, having symptoms at rest caused reduced QOL more than exertional symptoms that were relieved with rest. Fatigue was both the most common symptom and had the greatest impact on QOL. Reduction in sleep – often caused by worries over HVD – also reduced QOL.

Symptoms of any sort could reduce QOL directly by inducing fear, particularly if symptom onset was unpredictable, and by contributing to whether the participant defined themselves as ‘well’ or ‘unwell’ overall. Symptom-induced fear also indirectly reduced QOL when it caused participants to self-impose limitations on their physical activity. Participants’ perception of their disease could impact QOL by changing their view of their future and own identity, and respondents also had varying perceptions of the severity of their HVD, in particular, the likelihood of it being fatal.

HVD had a social impact: physical limitations caused some participants to reduce their community engagement, leaving some feeling that they were a burden or had let people down. HVD also impacted participants’ ability to spend quality time with their families, though most participants found their families to be a source of joy and support. Whether participants could work (or do hobbies if retired) was important for QOL, although some participants still in paid work found their HVD symptoms made work overwhelming.

External stressors impacted on QOL: to be in financial stress or for it to be difficult to go to hospital for check-ups reduced QOL. HVD impacted this link; some participants with symptomatic HVD found it made them less able to cope with external stressors. Emotional status was strongly linked to QOL: participants who considered themselves to be ‘stoic’ or having a positive attitude reported better QOL, and participants who felt negative emotions or unable to control those emotions reported poorer QOL. Medications affected QOL: participants reported some being difficult to take or causing side effects. Finally, the attitude of clinical staff impacted QOL: some participants described clinical staff as very good, while other participants reported experiences with staff that caused distress or distrust.

## DISCUSSION

These interviews provide insight into QOL in people with HVD. As expected, QOL in HVD is strongly linked to physical limitation. However, HVD has a broad effect on QOL across all areas of life: it affects participants’ sense of identity; impacts their involvement in their communities and engagement with their families; comes with burdensome symptoms that, independent of their physical effect, can cause confusion and fear and can restrict their ability to work; affects emotional status; comes with medications and treatment options that can be painful; and involves meeting many clinical workers, some of whom promote QOL and others who impair it.

In regards to physical limitations, a person’s ability relative to their perceived ‘normal’ was more important than their objective ability. As reported by one participant, to have a “disability… or ailment means you can’t do what you’ve always done”, and losing ‘normality’ drastically reduced QOL. This was irrespective of objective ability: one participant felt “anything’s possible” since they were able to “go walking” more after their valve replacement, whereas a younger participant who was still very fit felt grief due to losing the ability to play sport at high-level competitions. The relationship between physical limitation and QOL has not been quantitatively assessed in this study, but if we consider the effect of physical limitation and QOL in heart failure (being the most common cause of mortality in patients with HVD), results vary: some studies found only moderate correlation between physical capacity and QOL,[5] while others found a stronger correlation as well as similar features to those found in this project, such as loss of prior roles, fear of exertion, and reduced social network.[6] A recent study found physical limitation was more important for QOL than metabolic markers (NT-proBNP) and echocardiographic imaging at rest.[7]

A key impact of symptoms was not the symptom itself but the worry and reduced activity it inspired. Exertional symptoms only appeared to have minimal effect on QOL. However, any symptom could inspire worry due to participants being aware that their diagnosis could potentially be fatal (even if the chance was low). Many participants, when experiencing any symptom, immediately worried about its significance. Research in patients with HF shows a similar finding: that symptoms reduce QOL directly but are also linked to depression, which indirectly reduces QOL.[8]

The relationship between VRR and QOL varied. Participants who recovered smoothly from VRR did well, particularly if they viewed their HVD as a mechanical issue which could be totally cured. However, participants whose recovery from VRR was longer and more painful than they expected had impaired QOL, as did those who had more remaining symptoms than expected. While some participants who had not received VRR looked forward to it, others found that a potential operation in the future inspired worry. This was regardless of whether the VRR would be by open surgical procedure or transcatheter: most participants regarded both simply as ‘an operation’. Clinicians viewed valve replacements as requiring careful assessment, and acknowledged that it could not always entirely cure HVD. Unlike native heart valve disease, changes in QOL after VRR has received significant attention from researchers. It has been found that poorer heart function before valve replacement is associated with lesser improvement in QOL after intervention.[9] Another study found poorer QOL after intervention was associated with depression and reduced physical capacity before intervention. That study also found ‘belief in intervention’ to be predictive of QOL after intervention, as did our study, which found better QOL post-VRR if patients viewed VRR as something that could “fix” the (perceived as) mechanical problem of their HVD.

Expectations of the future had varying impact on QOL. Participants who had positive expectations of the future had the best QOL, but participants who had positive expectations then experienced poorer outcomes than expected had a reduced QOL. However, the worst QOL was in those with negative expectations of the future. This has been found in HF cohorts; people who expected deterioration in future had poorer QOL.[10]

Finally, we discuss the impact of fatigue on QOL, both for its impact on QOL but also to highlight the discrepancy between the clinical approach to fatigue and its impact on people with HVD. Fatigue was not only the most common symptom experienced, but also had the most devastating impact on QOL. A person with symptoms on exertion starts activity and then has to stop; a person with fatigue cannot start activity at all. While few participants had severe SOB or chest pain, many had severe fatigue. Even those experiencing milder fatigue often reported it was their most burdensome symptom, as it impaired their ability to do activities they considered important. In short, many participants felt their fatigue had not simply reduced their QOL but had taken their lives away from them. Fatigue is a common symptom in HF, and is strongly associated with QOL as well as mortality and is therefore an important clinical consideration.[11] Even without having developed acute HF, patients with symptoms of HF have increased fatigue and decreased QOL.[12]

However, rather than asking about fatigue, participants reported that their clinicians asked only about chest pain and SOB. This lack of investigation by clinicians into patients’ fatigue is not surprising, as fatigue is a vague and nonspecific symptom. Fatigue can be the primary complaint in anything from renal or liver disease to depression; it is the most common presenting complaint in people with liver disease, affecting 53% of those with chronic hepatitis,[13] and occurs in 70-97% of those with renal disease.[14] Therefore, whether fatigue would be useful to determine management strategies, and in particular valvular intervention, requires further research.

### Limitations

Many individuals with HVD had other comorbidities. As a result, findings of this study may be contributed to by comorbidities rather than just features of HVD. Nevertheless, HVD rarely exists in isolation; it is one part of an individual’s holistic health. To exclude all comorbidity would exclude real experiences of this cohort. Additionally, small sample sizes prevented comparison of symptoms and of QOL between groups of varying comorbidity and severity of valve lesions.

This project was limited by COVID-19, as nationwide lockdowns prevented the use of in person focus groups; the HVD cohort in New Zealand is often elderly, and many participants did not feel comfortable with navigating a digital group discussion. Focus groups were therefore not included in the study, and individual interviews used only. Focus groups, however, can elucidate perspectives that an interviewer without the disease state of interest cannot, and so the potential data here was lost.

Finally, this project was conducted in southern New Zealand, in a sample of mostly New Zealand European ethnicity. It is unclear if these findings would apply in a cohort of patients in other circumstances. In particular the Indigenous population in the Northern Territory of Australia is likely to face different challenges to NZ Europeans or Māori with HVD in New Zealand; a similar study conducted in this population may come to different conclusions about contributors to QOL.

## CONCLUSIONS

HVD has a major impact on all aspects of QOL. Although physical limitations play a significant role, many other aspects of QOL are also affected, such as sense of identity, engagement with community and family, and work. Fatigue is central to QOL in patients with HVD, but is rarely assessed by healthcare workers. Future research should examine how fatigue changes after procedural intervention, and whether it is specific enough to warrant identification as a symptom prompting procedural intervention.

## Data Availability

All data produced in the present study are available upon reasonable request to the authors

## ACKNOWLEDGEMENTS

This study was funded by the Department of Medicine, University of Otago, New Zealand. No authors had competing interests. A.P.: Conceptualization, Data curation; Formal analysis, Investigation, Methodology, Project Administration, Writing - original draft; G.W.: Conceptualization, Formal analysis, Methodology, Supervision, Writing - review and editing; R.W.: Validation, Writing - review and editing; P.B.: Methodology, Data curation, Validation, Writing - review and editing; R.S.: Validation, Writing - review and editing; P.A.: Validation, Writing - review and editing; R.R.T: Validation, Writing - review and editing; C.J.: Conceptualization, Methodology, Writing - review and editing; D.M.: Conceptualization, Methodology, Writing - review and editing; S.C.: Conceptualization, Formal analysis, Funding Acquisition, Investigation, Methodology, Project Administration, Resources, Supervision, Writing - review and editing;

## APPENDIX

**Appendix Table One:** Interview Structure.

| Initial structure<br>(Determined by first four interviews) | Final structure |
| --- | --- |
| Participant's definition of QOL/wellbeing | Participant's definition of QOL/wellbeing |
| What a 'good' and a 'bad' day consisted of | Participant's current QOL |
| How a 'bad' day could become a 'good' one | Progression of QOL over time |
| Checklist of items volunteered by other participants as important without the interviewer specifically asking, and whether the current participant thought they were important: Family, physical independence, pain, and challenges. | Checklist of items volunteered by other participants: Fatigue, pain playing a defining role in life, medications, and experiences with doctors and the healthcare system |
| Effect of HVD on QOL. If no significant effects had been noted to this point in the interview, the participant was asked to go through a typical day, and note the impacts of HVD at each point. |  |
| Progression of QOL over time |  |

*HVD = Heart Valve Disease; QOL = Quality of Life*

**Appendix Table Two:** Physical Factors and Quality of Life.

| Theme | Sub-themes | Example quotes / summaries |
| --- | --- | --- |
| Physical limitations | Able to do 'normal' activities | • QOL defined as "being able to do your usual activities", and "if someone can live comfortably the way they like to, with no problems going about their daily issues" |
|  | Life changed due to limitations | • Feeling "trapped" by HVD |
|  | Loss of physical independence, +/- embarrassment | • "would hate to be dependent on someone"<br>• "When you can't do certain things, like walk to the bathroom...that then becomes associated with this really low quality of life" |
|  | Loss of identity | • "limitations in one area; that reminds you of what you once were capable of and are no longer. I think that can evoke regret, and a bit of grief in terms of what once was"<br>• Participant was a high-achieving person prior to HVD but had to step down from roles, and has difficulty with even small tasks |
|  | Physical burden | • Walking even a short distance feels like "walking through porridge" or a "swamp" |
|  | Loss of social activity | • Sometimes have trouble keeping up with friends on group trips |
|  | Demotivation | • "I'm not really on top of these things enough to take it as it comes"<br>• "I think a reasonable amount of stoicism would sum up my character" |
| Symptoms: | Worst QOL symptoms at rest | • Worst QOL symptoms at rest |
|  | Pain worse than expected; felt uninformed | • "I definitely wasn't fully informed about the amount of pain included [for VRR]" |
| Symptoms: Fatigue | Fatigue has largest impact on QOL | • QOL defined as 'Having the energy to do things'<br>• Being unable to do basic tasks due to fatigue |
|  | Fatigue can be very severe but also nonspecific | • Fatigue is almost constant – becomes 'usual' |
| Sleep | Good sleep helps QOL; poor sleep reduces it | • Kept awake by worries over HVD or by palpitations |
| Work | Able to vs unable to work | • QOL defined as "being able to work"<br>• Has worked very hard all of life; work is part of identity |
|  | Overworked | • Having to take things slowly due to HVD |
| Situation | Difficulties accessing healthcare | • Biggest impact of HVD was hospital visits disrupting life |
|  | External circumstances | • Too high-risk for intervention = poorest QOL<br>• Being able to afford a comfortable lifestyle helps QOL |
|  | HVD makes dealing with external circumstances harder | • "[HVD was] only a part...Just the straw that broke the camel's back." |
| Medications and/or Treatment | Medications burdensome vs easy to take | • Feeling emotionally flat due to medication vs. medication having no side effects<br>• Medications can be annoying to take if there are specifications on when to take them and what to have them with<br>• Injections for RHD are painful |
|  | Painful recovery from VRR | • Recovery from surgical VRR is painful |

HVD = Heart Valve Disease; QOL = Quality of Life; VRR = Valve Replacement

**Table Three:**
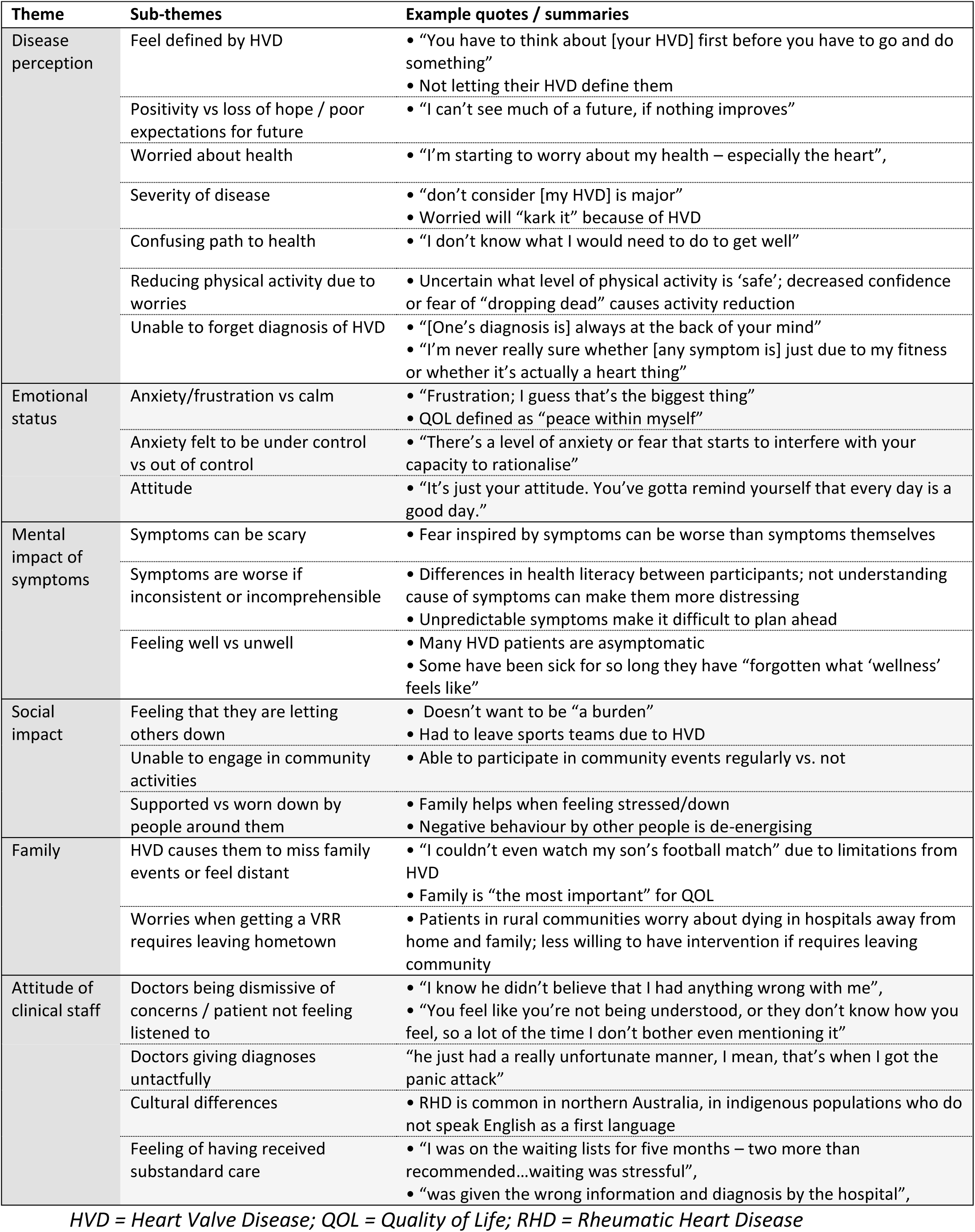
Perspective, Stressors, and Quality of Life.

| Theme | Sub-themes | Example quotes / summaries |
| --- | --- | --- |
| Disease perception | Feel defined by HVD | • “You have to think about [your HVD] first before you have to go and do something”<br>• Not letting their HVD define them |
|  | Positivity vs loss of hope / poor expectations for future | • “I can’t see much of a future, if nothing improves” |
|  | Worried about health | • “I’m starting to worry about my health – especially the heart”, |
|  | Severity of disease | • “don’t consider [my HVD] is major”<br>• Worried will “kark it” because of HVD |
|  | Confusing path to health | • “I don’t know what I would need to do to get well” |
|  | Reducing physical activity due to worries | • Uncertain what level of physical activity is ‘safe’; decreased confidence or fear of “dropping dead” causes activity reduction |
|  | Unable to forget diagnosis of HVD | • “[One’s diagnosis is] always at the back of your mind”<br>• “I’m never really sure whether [any symptom is] just due to my fitness or whether it’s actually a heart thing” |
| Emotional status | Anxiety/frustration vs calm | • “Frustration; I guess that’s the biggest thing”<br>• QOL defined as “peace within myself” |
|  | Anxiety felt to be under control vs out of control | • “There’s a level of anxiety or fear that starts to interfere with your capacity to rationalise” |
|  | Attitude | • “It’s just your attitude. You’ve gotta remind yourself that every day is a good day.” |
| Mental impact of symptoms | Symptoms can be scary | • Fear inspired by symptoms can be worse than symptoms themselves |
|  | Symptoms are worse if inconsistent or incomprehensible | • Differences in health literacy between participants; not understanding cause of symptoms can make them more distressing<br>• Unpredictable symptoms make it difficult to plan ahead |
|  | Feeling well vs unwell | • Many HVD patients are asymptomatic<br>• Some have been sick for so long they have “forgotten what ‘wellness’ feels like” |
| Social impact | Feeling that they are letting others down | • Doesn’t want to be “a burden”<br>• Had to leave sports teams due to HVD |
|  | Unable to engage in community activities | • Able to participate in community events regularly vs. not |
|  | Supported vs worn down by people around them | • Family helps when feeling stressed/down<br>• Negative behaviour by other people is de-energising |
| Family | HVD causes them to miss family events or feel distant | • “I couldn’t even watch my son’s football match” due to limitations from HVD<br>• Family is “the most important” for QOL |
|  | Worries when getting a VRR requires leaving hometown | • Patients in rural communities worry about dying in hospitals away from home and family; less willing to have intervention if requires leaving community |
| Attitude of clinical staff | Doctors being dismissive of concerns / patient not feeling listened to | • “I know he didn’t believe that I had anything wrong with me”,<br>• “You feel like you’re not being understood, or they don’t know how you feel, so a lot of the time I don’t bother even mentioning it” |
|  | Doctors giving diagnoses untactfully | • “he just had a really unfortunate manner, I mean, that’s when I got the panic attack” |
|  | Cultural differences | • RHD is common in northern Australia, in indigenous populations who do not speak English as a first language |
|  | Feeling of having received substandard care | • “I was on the waiting lists for five months – two more than recommended...waiting was stressful”,<br>• “was given the wrong information and diagnosis by the hospital”, |

HVD = Heart Valve Disease; QOL = Quality of Life; RHD = Rheumatic Heart Disease

## Notes

### Competing Interest Statement

The authors have declared no competing interest.

### Author Declarations

The Health and Disabilities Ethics Committee, New Zealand, gave ethical approval for this work (HDEC, ethics reference 19/NTA/163). All individuals gave informed consent to take part in the study, and participant anonymity was preserved in analysis and presentation.

